# Robust cancer crowdfunding predictions: Leveraging large language models and machine learning for success analysis

**DOI:** 10.1101/2025.02.24.25322692

**Authors:** Abhishikta Roy, Vineet Srivastava, Lokesh Boggavarapu, Ranganathan Chandrasekharan, Edward Mensah, John Galvin, Runa Bhaumik

## Abstract

**Background:** In the field of medical crowdfunding prediction, traditional statistical methods have long been the standard. Machine learning algorithms are popular because they can model complex relationships between variables, capture interactions, and provide more accurate predictions, even when input variables are highly correlated. Furthermore, previous research has largely overlooked the quantitative assessment of success levels and the selection of key predictors. To address these limitations, a novel approach is needed that leverages advanced machine learning techniques.

**Objective:** This study aimed to address these gaps by proposing a robust feature engineering approach that leverages the capabilities of large language models (LLMs). The goal was to extract the success determinants using a large language model for a cancer crowdfunding campaign. Furthermore, this study evaluated the performance of four machine learning algorithms in predicting campaign success and quantitatively assessed the level of success.

**Method:** We separately analyzed linguistic and social determinants of health features to understand how much each factor contributes to a crowdfunding campaign’s success. These features were generated using a large language model (GPT-4o). A random forest algorithm with a permutation technique was used to rank the features. We comparatively evaluated the prediction accuracy, sensitivity, and specificity of four machine learning algorithms, random forest, gradient boosting, logistic, and elastic net, using a 10-fold cross-validation.

**Results:** Gradient Boosting consistently outperforms the other algorithms in terms of sensitivity (consistently around 0.786 to 0.798), indicating its superior ability to identify successful crowdfunding campaigns using linguistic and social determinants of health features. The permutation importance score reveals that for severe medical conditions, income loss, chemotherapy treatment, clear and effective communication, cognitive understanding, family involvement, empathy and social behaviors play an important role in the success of campaigns.

**Conclusions:** This study highlights the critical role of linguistic, social, demographic, and medical features in predicting the success of cancer crowdfunding campaigns, with risk communication and medical severity emerging as key predictors. The study also suggests the need for more nuanced and optimized models and improved income protection and healthcare policies to reduce reliance on crowdfunding for cancer treatment.

## Introduction

The cost of cancer care in the US is increasing rapidly due to various factors, including technological advancements, expensive cutting-edge therapies, and improved access to treatment. Cancer patients and their families often face substantial financial consequences such as borrowing money, spending less on food, going into debt, and/or declaring bankruptcy ^1–3^. These issues are exacerbated by a lack of medical insurance or lack of coverage due to healthcare regulations such as the Affordable Care Act. As a result, patients and caregivers are seeking help from other sources to raise funds for medical care ^2^.

Crowdfunding has become a significant tool for raising money through social media and web-based platforms like Indiegogo, Kickstarter, and GoFundMe generating $34.4 billion in 2015 alone ^4–7^. Medical crowdfunding constitutes a significant portion of these campaigns in the United States due to gaps in insurance coverage and the prohibitive costs of medical treatments, such as cancer care, even for those with insurance ^8,9^. Success factors for these campaigns include demographic attributes, racial background, campaign images, and textual features, social media characteristics, with campaigns for children attracting more donations than those for adults ^6,7,9,10^. In cancer treatment crowdfunding, 65.4% of campaigns involve advanced-stage patients, with detailed information about cancer types, treatments, and costs leading to higher fundraising success ^11–15^. Additionally, research has characterized the use of crowdfunding to support oncology care needs, examining associations between insurance status and other characteristics related to unmet financial obligations ^13^.

The field of crowdfunding prediction has primarily relied on traditional statistical methods, such as linear and logistic regression, in prior studies. These methods assume that the input variables are independent from each other. However, when there are correlations among the input variables, using regression methods can lead to larger prediction errors. To overcome this limitation, researchers have turned to machine learning techniques. Machine learning algorithms have proven to be effective in analyzing hidden associations within large datasets and identifying complex patterns ^16,17^. As a result, researchers utilized machine learning algorithms to predict the success of crowdfunding campaigns ^18–20^. This shift in methodology allows for better prediction accuracy and the discovery of more nuanced relationships among the input variables. For instance, machine learning algorithms like Support Vector Machine (SVM), Decision Trees (DT), and K-nearest neighbor (KNN) have been utilized to predict the success of projects ^18,19,21–24^. Additionally, algorithms such as XgBoost, Gradient Boosting, Random Forest, and Generalized Linear Model (GLM) have been used to construct prediction models ^25^.

Previous crowdfunding research has demonstrated that machine learning methods can effectively model complex relationships between variables, capture interactions, and yield more accurate and insightful predictions. However, there has been a lack of thorough analysis regarding the selection of predictors and campaign success measures. Furthermore, the application of large language models in the crowdfunding domain remains relatively unexplored. The only study^26^ to date has generated insights on medical financial hardship patterns and unmet social needs by leveraging generative AI on GoFundMe cancer crowdfunding campaigns using logistic regression.

In this study, we explored the use of large language and machine learning models to generate and identify predictors for successful campaigns. The large language model is particularly helpful in capturing the semantic meaning of words and phrases in internet-based texts by using word embeddings and contextual embeddings, which were trained on large, diverse texts. GPT-4o ^27^, developed by OpenAI, is a large language model known for its conversational text generation capabilities. We utilized GPT-4o and prompted relevant campaign information to extract and analyze linguistic and social determinants of health (SDOH) factors. We also provided a robust feature selection based on the permutation technique for a better understanding of the model.

Our research has three primary objectives:

1. Leverage large language models like GPT-4o to generate an wide range of features.
2. Implement a robust feature selection strategy to explain the contribution of each feature.
3. Conduct a comprehensive assessment of campaign success metrics and potential predictors leveraging an ensemble approach.

To the best of our knowledge, this study is the first to use a machine-learning algorithm to establish an effective prediction model for GoFundMe cancer crowdfunding campaigns with a robust feature selection method. By integrating innovative machine learning algorithms and feature selection strategies into crowdfunding research, we not only advance the academic understanding of this domain but also provide actionable insights that can be directly applied by policymakers and campaign organizers. These advancements are essential for addressing the growing reliance on crowdfunding in the healthcare sector and ensuring that support reaches those who need it most.

## Methods

### Data

The data for this study was collected from the GoFundMe crowdfunding platform, which is the world’s largest platform in terms of both the total amount of funding raised and the total number of active campaigns. Founded in 2010, GoFundMe features campaigns for various categories, including medical, memorials, emergencies, and charitable causes. The data for this analysis was extracted using a combination of API access and web scraping techniques. GoFundMe stores its data in Algolia, a search and analytics engine. We obtained the necessary API Key, Secret Key, and Index name by inspecting the network tab on the GoFundMe website. We connected to the Algolia API using a Python program and retrieved the data. The API provided access to all records stored in the specified index, which we then processed and stored for further analysis. The data consists of 4990 campaigns regarding a patient’s oncologic treatment and personal and financial life that began between January 3rd, 2023, and December 31st, 2023, with funding targets between US$800 and US$1000,000.

### Success Outcome

A campaign can be considered successful if it raises a significant percentage of its goal (e.g., 80% or 100%). Campaigns that reach or exceed their fundraising goal would be considered highly successful. success ratio, defined as the ratio of funds raised to the campaign goal, was converted to a binary variable to evaluate different success thresholds.

### Predictors of Campaign Success

Building on previous studies, we identified and added new predictors for success in medical crowdfunding campaigns on the GoFundMe site, which are described below and listed in Tables 1 and 2 of the supplementary file.

### Crawled Features

We collected key campaign data, including launch date, title, description, current raised amount, target goal, number of donors, and campaign duration. These details provide insights into each campaign’s progress and engagement.

### Linguistic Features

Language and psychological measures significantly influence crowdfunding success. We utilized the LIWC internal dictionary (Pennebaker et al., 2015) to analyze features such as analytical thinking, clout, authenticity, tone, cognition, politeness, empathy, social behavior, lifestyle, religion, and temporal aspects (past, present, future tense). These linguistic features help understand the multifaceted aspects contributing to a campaign’s success.

### Demographic and Social Factors

Demographic information includes the age group and gender of the beneficiary or campaigner. We assessed various health and social status factors, including cancer site, stage, therapy type, diagnosis date, treatment status, and associations with healthcare organizations.

### Emotional States

The mental and emotional state of individuals was assessed for stress, anxiety, behavioral dysfunctions, chronic diseases, and sentiment (positive, negative, neutral). The trust level in medical care facilities was also evaluated.

### Employment and Education Factors

We considered employment status, work disruption, school absenteeism, and financial impacts such as income loss and struggles with medical, housing, food, and transportation expenses.

### Medical Procedures and Treatment Factors

This includes laboratory procedures, diagnostic procedures, specific cancer treatments, preventive procedures, alternative therapies, and mental health treatments.

### Normalization

We applied min-max normalization to variables with large value ranges (target amount, raised amount, number of donors, and campaign duration) to minimize their impact on prediction results. GPT-4o was used to standardize measures of linguistics and campaign predictors within 0 to 1. The predictors for social and demographic identity, as well as medical-related, were extracted as a binary (yes/no) and were dummy-coded.

## Methods

### OpenAI (GPT4o)

It is a highly advanced language model capable of generating human-like text. GPT-4o has been trained on a large corpus of data and is known for its ability to understand context and produce coherent and contextually relevant responses.

### Logistic Regression with Elastic Net Regularization

Logistic Regression is a linear model used for binary classification problems. It models the probability that a given input belongs to a particular class. The logistic function (sigmoid function) is used to map predicted values to probabilities. Logistic Regression with Elastic Net Regularization ^16^ combines the properties of both L1 (Lasso) and L2 (Ridge) regularization techniques. This approach adds a penalty to the model based on the magnitude of the coefficients, helping to prevent overfitting and manage multicollinearity among features. It is computationally less intensive and faster to train compared to ensemble methods. However, it assumes a linear relationship between the features and the log-odds of the outcome, which may not capture complex patterns.

### Random Forest

Random Forest is an ensemble learning method ^17^ that constructs multiple decision trees during training and outputs the class that is the mode of the classes (for classification) or mean prediction (for regression) of the individual trees. It improves the model’s accuracy and robustness by reducing overfitting, which is a common problem with individual decision trees.

### Gradient Boosting

Gradient Boosting is another powerful ensemble technique^28^ that builds models sequentially. Each new model attempts to correct the errors made by the previous models. Unlike Random Forest, which builds trees in parallel, Gradient Boosting builds trees one at a time, where each new tree helps to correct the errors of the previous one.

### Feature Importance

Tree-based models, like Random Forests and Gradient Boosting, inherently capture interaction terms and non-linear relationships due to their structure. They split the data based on feature values, effectively capturing interactions and non-linearities without the need for explicit feature engineering. The permutation technique is used to calculate the importance of each feature. This method shuffles each feature individually and measures the change in the model’s performance to determine the importance of that feature.

### Experimental Setup

We conducted two sets of comparative experiments based on machine learning algorithms to predict the success of crowdfunding projects. We selected campaign and linguistic features as the independent variables in the first experiment. In the first experimental setup, we used a random forest algorithm with a default parameter setting in Scikit Learn Library in Python for parameter selection to rank the linguistic features based on their importance or relevance in predicting crowdfunding success measures. The reason for using default parameters is that the grid search parameter optimization technique did not result in improved model performance on the training data compared to the default settings. We used the permutation importance technique to determine the significance of each feature in a predictive model.

Unlike traditional feature importance measures in Random Forests, which are based on the reduction in impurity (like Gini impurity or entropy) from splits in the trees, permutation importance evaluates the impact of each feature on the model’s performance by shuffling its values. We evaluated the four learning algorithms mentioned above and compared the prediction performances. We repeated the same procedure on social, demographic, and medical-related features in the second experiment. The data was split into training and testing sets of a 70/30 % ratio. All machine learning model performances were validated using 10-fold cross-validation on training data. The testing set was used to validate our random forest model during feature selection. We used sensitivity, specificity, and accuracy for our evaluation metrics. Sensitivity was calculated by dividing the number of true positives (campaign success) by the sum of true positives and false negatives. Specificity was calculated by dividing the number of true negatives (unsuccessful campaigns) by the sum of true negatives and false positives. Accuracy was calculated by dividing the sum of true positives and true negatives by the total population.

## Results

### Descriptive Analysis

In our analysis, we used a dataset of crowdfunding campaigns to explore their success in relation to their progress towards the established goal amounts. The descriptive statistics of the raised amount provide valuable insights into the overall distribution and characteristics of the dataset. Among the 4984 campaigns analyzed, the mean raised amount is $ 20016.67 with a median of $ 11819.0. Similarly, the mean for the goal amount amongst the scraped links is $ 40373.73, and the median is $ 25000.00. We divided the dataset into five distinct groups by categorizing the data into percentile ranges based on the distribution of a goal amount. Each group is of equal size: (799.999, 10000], (10000,20000], (20000, 30000], (30000, 50000], and (50000, 1000000]. Figures 1 and 2 depict the distribution of the total number of campaigners and the total number of donors in each bucket.

**Figure 1.**
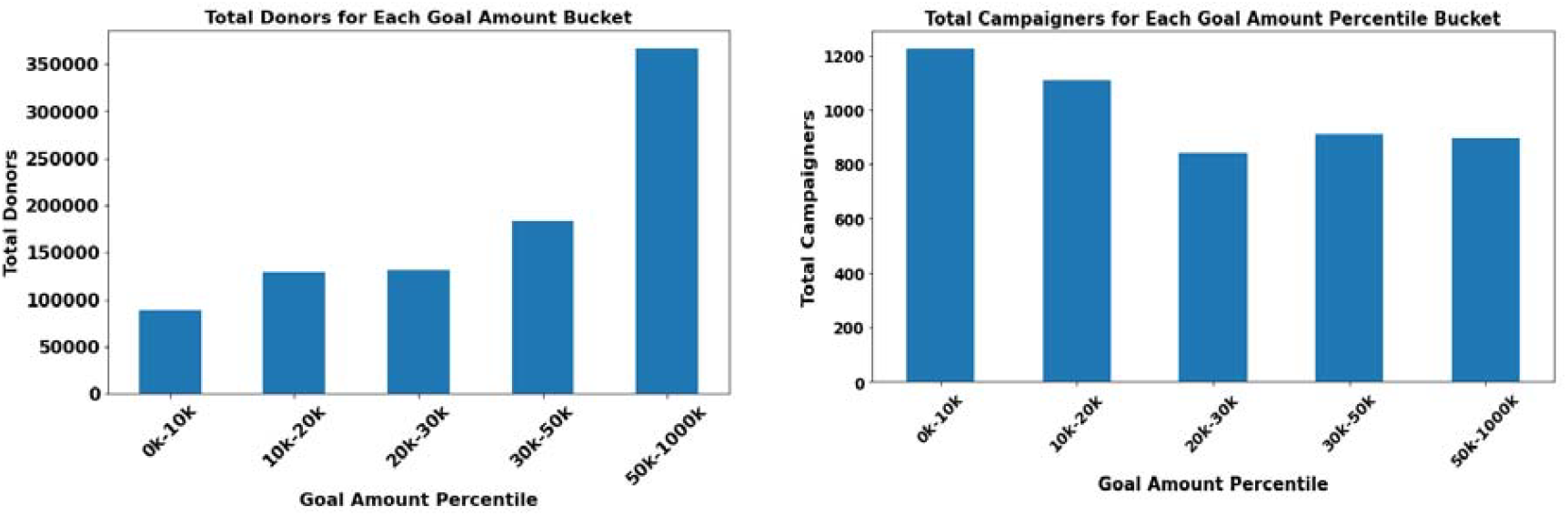
Distribution of the total number of donors and campaign in each Goal Amount bucket

Figure 1 presents an analysis of crowdfunding campaigns based on goal amounts and their relationship with the total number of donors and campaigners. Higher goal amounts are associated with a significantly higher number of donors. Campaigns with goals between $50,000 and $1000,000 attract the most donors. This trend suggests that higher goals may signal more substantial and potentially more impactful projects, attracting more backers. Lower goal amounts ($799.999 to $10,000) have the highest number of campaigns, indicating that many campaigners prefer setting lower, more achievable goals. As the goal amount increases, the number of campaigns decreases, suggesting that fewer campaigners aim for higher funding targets. These two figures provide valuable insights for crowdfunding campaigners. Setting a higher goal amount can potentially attract more donors and maximize success.

**Figure 2:**
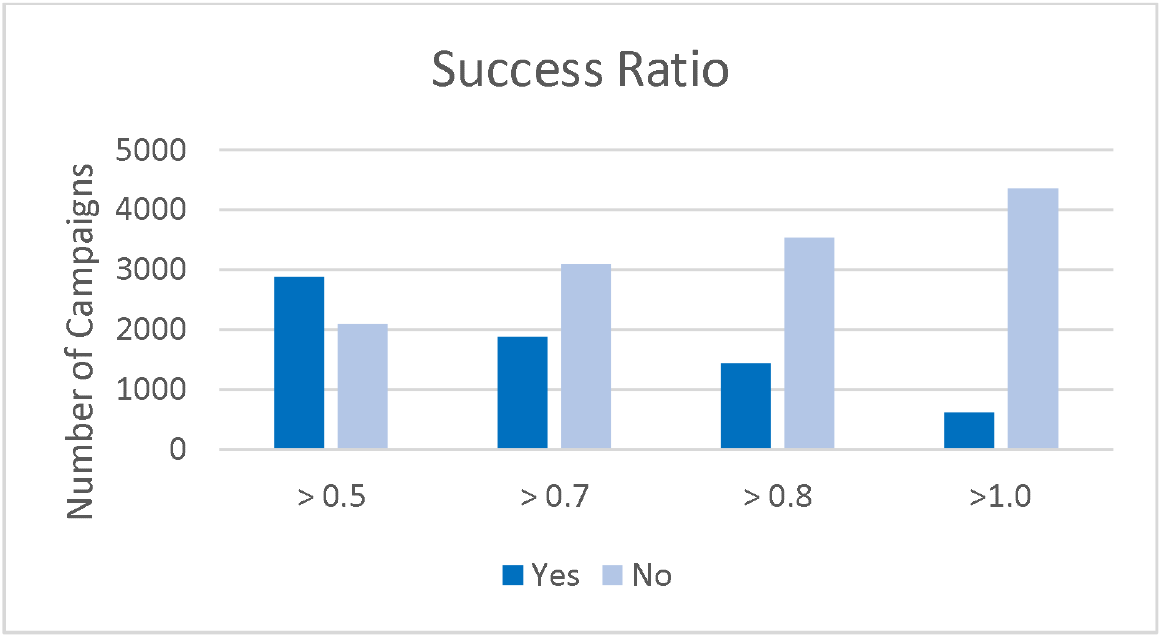
Distribution of crowdfunding campaigns based on their success ratio

As the success ratio threshold increases, the proportion of successful campaigns decreases, while the number of unsuccessful campaigns increases. This trend is expected, as higher thresholds represent more ambitious funding goals, which are harder to achieve.

**Figure 3.**
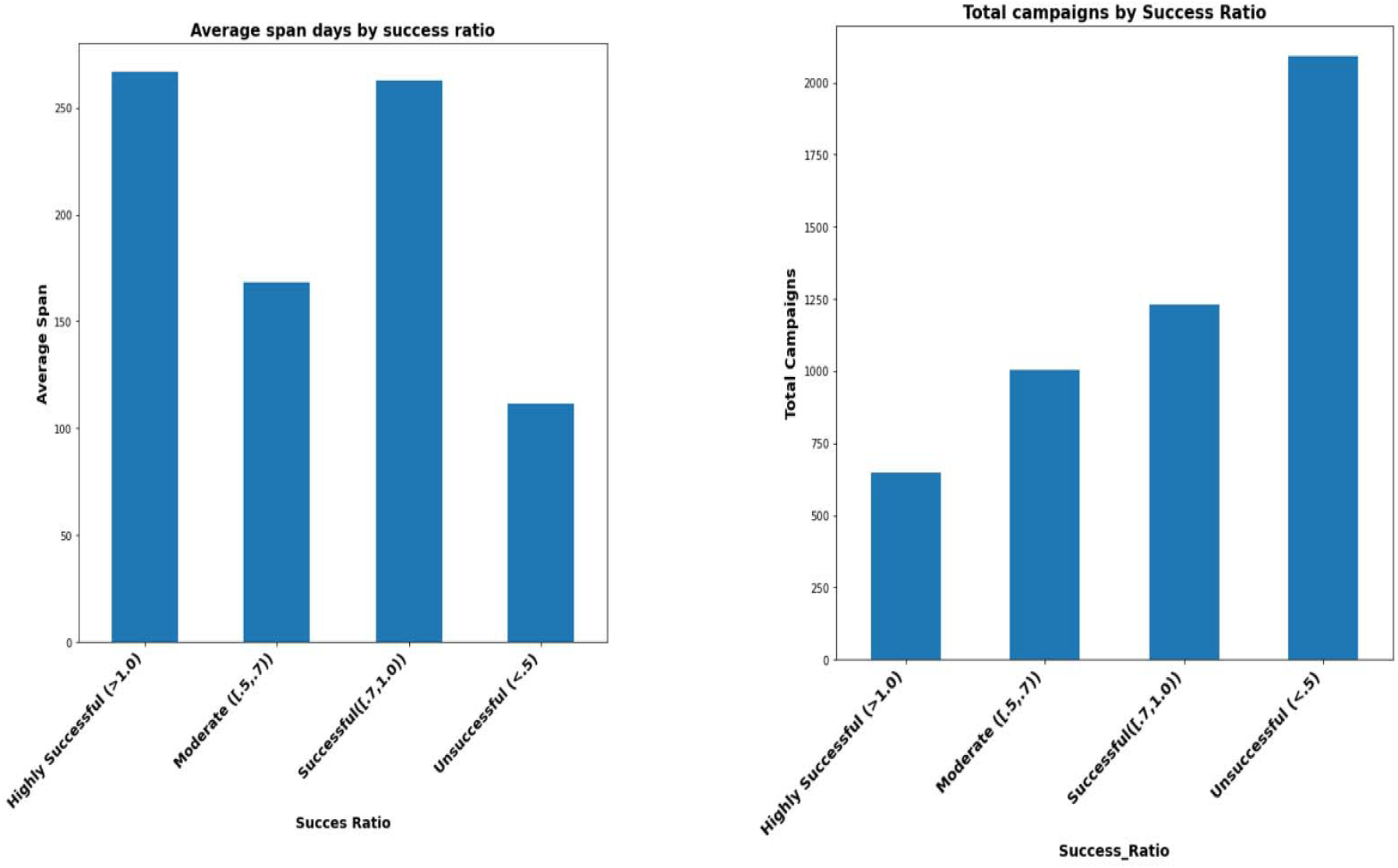
Distribution of span and campaign by success measures

The charts analyze crowdfunding campaigns based on their success ratios, examining the average span of the campaigns and the total number of campaigns in each success category. Campaigns in the Highly Successful category have the longest average span, slightly over 250 days. It indicates that highly successful campaigns tend to have a longer duration.

### Performance Evaluation of Machine Learning Models

**Table 1:** Performance Evaluation of ML Algorithms at a 0.7 Success Threshold for SDOH Factors.

|  | Random Forest |  |  | Gradient Boosting |  |  | Logistic |  |  | Elastic Net |  |  |
| --- | --- | --- | --- | --- | --- | --- | --- | --- | --- | --- | --- | --- |
|  | Acc | Sen | Spec | Acc | Sen | Spec | Acc | Sen | Spec | Acc | Sen | Spec |
| **top3** | 0.770 | 0.660 | 0.830 | 0.790 | 0.690 | 0.838 | 0.637 | 0.013 | 0.999 | 0.648 | 0.043 | 0.998 |
| **top6** | 0.770 | 0.660 | 0.830 | 0.780 | 0.690 | 0.835 | 0.637 | 0.013 | 0.999 | 0.649 | 0.046 | 0.998 |
| **top9** | 0.760 | 0.650 | 0.830 | 0.780 | 0.690 | 0.838 | 0.637 | 0.014 | 0.999 | 0.649 | 0.047 | 0.998 |
| **top12** | 0.770 | 0.680 | 0.840 | 0.790 | 0.690 | 0.838 | 0.639 | 0.018 | 0.999 | 0.651 | 0.052 | 0.998 |
| **top15** | 0.760 | 0.610 | 0.846 | 0.790 | 0.690 | 0.842 | 0.639 | 0.020 | 0.999 | 0.651 | 0.052 | 0.998 |
| **top18** | 0.770 | 0.630 | 0.853 | 0.790 | 0.69 | 0.839 | 0.640 | 0.022 | 0.999 | 0.651 | 0.052 | 0.998 |
| **top21** | 0.770 | 0.610 | 0.856 | 0.790 | 0.695 | 0.840 | 0.640 | 0.022 | 0.999 | 0.650 | 0.052 | 0.997 |
| **top24** | 0.760 | 0.580 | 0.859 | 0.790 | 0.693 | 0.843 | 0.640 | 0.022 | 0.999 | 0.651 | 0.053 | 0.997 |
| **top28** | 0.770 | 0.610 | 0.863 | 0.790 | 0.693 | 0.843 | 0.640 | 0.022 | 0.999 | 0.650 | 0.052 | 0.997 |

Table 1 presents the performance evaluation of four machine learning (ML) algorithms—Random Forest, Gradient Boosting, Logistic Regression, and Elastic Net—at a 0.7 success threshold, specifically for Social Determinants of Health (SDOH) factors. The reported performance metrics include accuracy (Acc), sensitivity (Sen), and specificity (Spec) across different subsets of top features (ranging from top 3 to top 28). Gradient Boosting consistently outperforms the other algorithms in terms of sensitivity (consistently around 0.786 to 0.788), indicating its superior ability to identify successful crowdfunding campaigns. Random Forest offers a balanced performance with good accuracy (.767 to .769) and specificity (.831 to .863), making it a reliable choice for predicting crowdfunding success while capturing non-linear relationships and interactions. Logistic Regression and Elastic Net exhibit strong specificity (.997 to .999) but poor sensitivity (.013 to .052), which suggests they may be overly conservative and are not able to predict successful campaigns correctly.

Based on the evaluation when using Gradient Boosting, selecting the top 3 to 12 features seems optimal. The top 3 features provide a strong balance of high accuracy and sensitivity with fewer variables, making the model simpler and potentially more generalizable. If one aims to maximize model performance with a bit more complexity, one can consider using up to the top 12 features, which slightly enhances specificity without sacrificing other metrics.

**Table 2:** Performance Evaluation of ML Algorithms at a 0.7 Success Threshold for Linguistic Factors.

|  |  | Random Forest |  |  | Gradient Boosting |  |  | Logistic |  |  | Elastic Net |  |
| --- | --- | --- | --- | --- | --- | --- | --- | --- | --- | --- | --- | --- |
|  | Acc | Sen | Spec | Acc | Sen | Spec | Acc | Sen | Spec | Acc | Sen | Spec |
| top3 | 0.766 | 0.686 | 0.815 | 0.798 | 0.723 | 0.843 | 0.631 | 0.025 | 0.998 | 0.648 | 0.076 | 0.996 |
| top6 | 0.782 | 0.691 | 0.837 | 0.797 | 0.723 | 0.842 | 0.634 | 0.034 | 0.998 | 0.65 | 0.081 | 0.995 |
| top9 | 0.785 | 0.688 | 0.844 | 0.797 | 0.721 | 0.842 | 0.635 | 0.036 | 0.998 | 0.654 | 0.092 | 0.995 |
| top12 | 0.794 | 0.694 | 0.855 | 0.799 | 0.725 | 0.843 | 0.643 | 0.062 | 0.995 | 0.659 | 0.113 | 0.991 |
| top15 | 0.786 | 0.667 | 0.859 | 0.798 | 0.727 | 0.84 | 0.643 | 0.062 | 0.995 | 0.659 | 0.112 | 0.992 |
| top18 | 0.788 | 0.666 | 0.862 | 0.797 | 0.722 | 0.842 | 0.643 | 0.066 | 0.994 | 0.66 | 0.12 | 0.988 |
| top21 | 0.788 | 0.654 | 0.869 | 0.793 | 0.718 | 0.839 | 0.643 | 0.066 | 0.994 | 0.66 | 0.119 | 0.988 |
| top24 | 0.782 | 0.634 | 0.873 | 0.797 | 0.723 | 0.842 | 0.645 | 0.073 | 0.992 | 0.662 | 0.128 | 0.987 |
| top28 | 0.788 | 0.647 | 0.874 | 0.796 | 0.723 | 0.84 | 0.644 | 0.074 | 0.991 | 0.66 | 0.122 | 0.987 |

The results indicate that careful feature selection is crucial for optimizing model performance for linguistic features. Gradient Boosting and Random Forest show that selecting around the top 12 features provides a strong balance between sensitivity and specificity, making them well-suited for predictive tasks in medical crowdfunding. Logistic Regression may require a different approach or fewer features to avoid unnecessary complexity, while Elastic Net’s performance suggests a moderate feature increase (up to 18-24) could be beneficial. Overall, the findings highlight the importance of tailored feature selection strategies to maximize the effectiveness of machine learning models in this domain.

### Campaign Success Determinants

**Figure 4.**
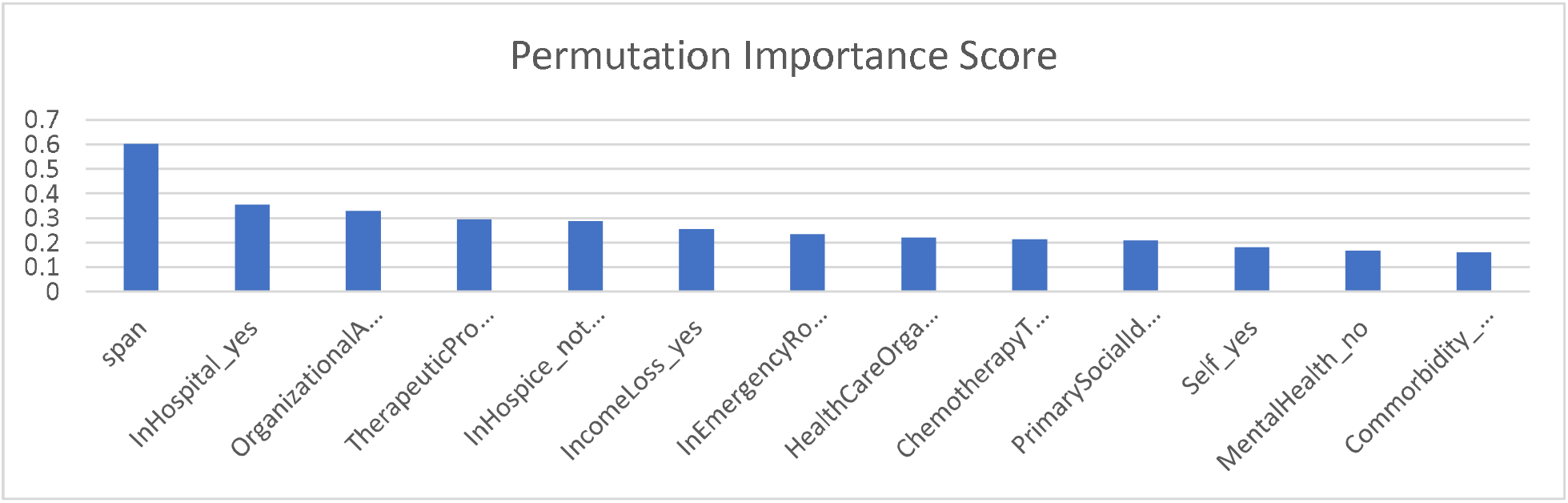
Campaign Success Factors for SDOH predictors

Figure 4 displays the top 13 important features (SDOH) in predicting success at a .7 success ratio for the Gradient Boosting algorithm. Permutation Importance score is a metric that indicates how much a particular feature contributes to the predictive power of a machine learning model. The number of donors and goal amount are the most significant predictors of success in the context of this medical crowdfunding model. To improve the visibility of importance scores for the remaining features, we have excluded these two variables from Figure 4 due to their large variation. The other features, while related to medical and demographic factors, have a much smaller impact on the model’s predictive accuracy. Based on the chart, The fact that being in the hospital or in unspecified hospice care is highly important highlights the severe medical conditions often associated with crowdfunding campaigns. Policymakers could use this information to identify areas where the medical system is failing to cover critical care, prompting discussions on improving hospital funding or hospice services. The importance of income loss suggests that financial hardship due to illness is a key driver of crowdfunding success. This implies that patients need better income protection policies or insurance solutions to support individuals who face financial difficulties due to medical conditions. The involvement of healthcare organizations as a feature suggests that campaigns supported by these organizations might have better access to resources than individuals. The importance of chemotherapy treatment as a feature suggests that crowdfunding campaigns associated with this treatment might attract more attention and support, possibly because donors recognize the high costs and seriousness of cancer treatment. Also, the female patient has a successful campaign.

**Figure 5.**
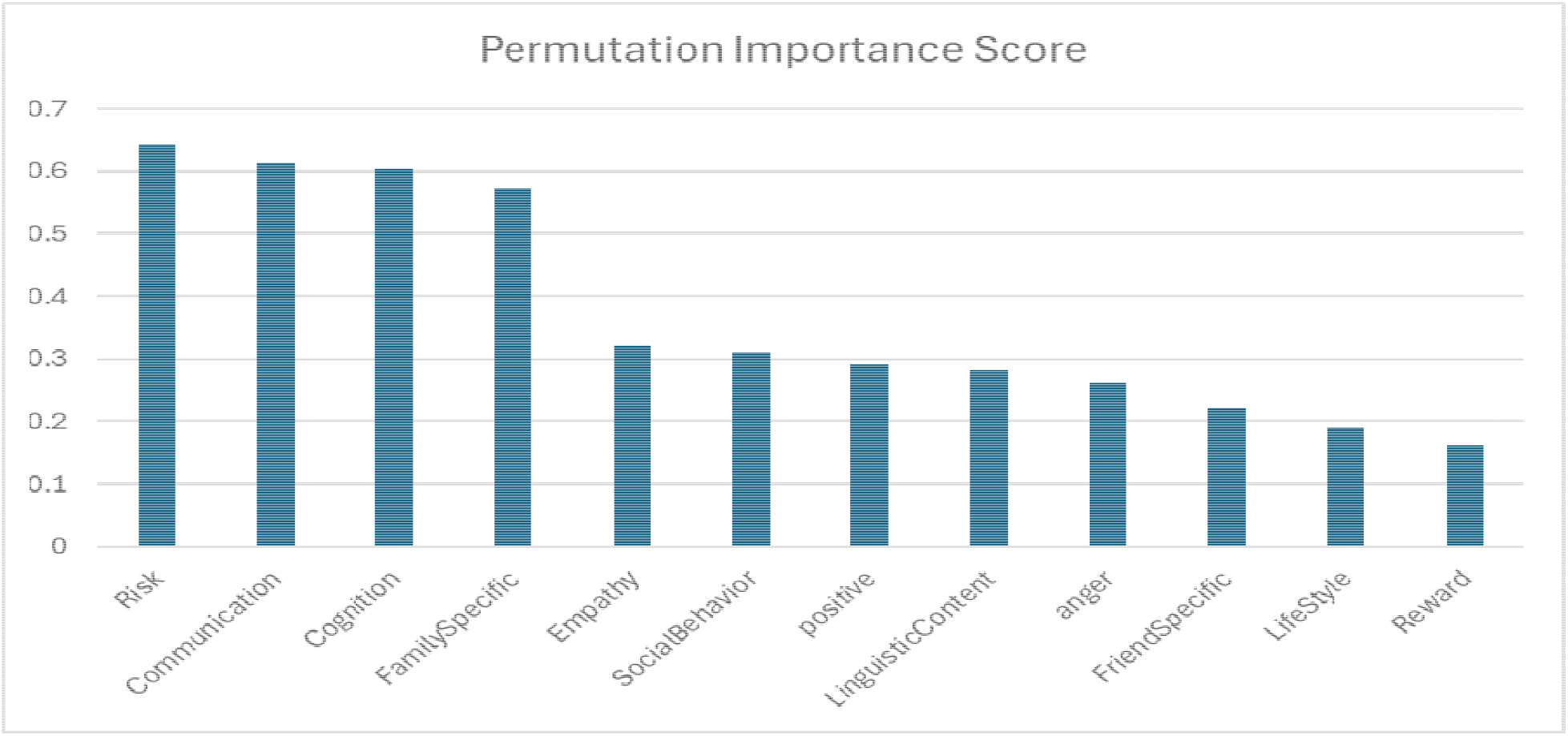
Campaign Success Factors for Linguistic Predictors

The chart displays the permutation importance scores of various linguistic features related to some aspect of behavior or decision-making, possibly in a medical or social context. The features are ranked by their importance in predicting an outcome, with higher scores indicating a greater influence on the model’s performance. Clear and effective communication, particularly around risk, while also fostering cognitive understanding and family involvement. Empathy and social behaviors should be cultivated in professional contexts, and positive messaging should be encouraged across various communication channels. Although less critical, addressing factors like anger, lifestyle, and reward could further enhance outcomes. These policy recommendations aim to leverage the most influential factors identified by the model to achieve better decision-making and outcomes in the relevant context.

## Discussion

### Principal Results

In this study, we conducted an empirical analysis of medical crowdfunding. The analysis of 4984 crowdfunding campaigns revealed key insights into the relationship between goal amounts, campaign success, and donor engagement. Campaigns with higher goals ($50,000-$100,000) attracted significantly more donors, indicating that ambitious targets may signal substantial projects, thus drawing greater support. Conversely, lower goal amounts ($799.99-$10,000) saw the highest number of campaigns, suggesting a preference for achievable targets among campaigners. Highly successful campaigns exhibited longer durations, averaging over 250 days. We also focused on deriving features using large language model and Feature selection using the Random Forest algorithm. We identified “donation_count_full,” “goal amount,” and “span” as critical predictors of success, with emotional and thematic features also contributing. Among the machine learning models evaluated, GB demonstrated superior performance (range: .786-.788) and True Positive Rate (range: .693-.696), outperforming RF, logistic, and EN. The analysis highlights the critical role of linguistic and Social Determinants of Health (SDOH) predictors in the success of crowdfunding campaigns. Linguistic features such as clear and effective communication, particularly regarding risk, along with positive messaging, empathy, and social behaviors, significantly influence campaign outcomes. SDOH predictors, including the severity of medical conditions (e.g., hospitalization or unspecified hospice care), income loss, and the involvement of healthcare organizations, are also key drivers of crowdfunding success. These findings suggest a need for improved hospital funding, better income protection policies, and stronger support from healthcare organizations. Additionally, campaigns associated with high-cost treatments like chemotherapy tend to attract more attention, underscoring the need for targeted support for patients undergoing such treatments.

### Limitations

This study has several limitations. The study primarily focuses on linguistic, social, demographic, and medical-related features. Other factors, such as donor engagement metrics or campaign visibility on social media platforms, have not been evaluated. While cross-validation helps in mitigating overfitting, the generalizability of the model to other types of crowdfunding campaigns beyond the specific dataset used (e.g., different medical conditions, geographic regions, or platforms) remains uncertain. The model’s performance may vary when applied to other datasets, limiting its broader applicability. The study relies on the available dataset for training and testing. If the dataset is not representative of the broader population of cancer crowdfunding campaigns (e.g., biased towards certain demographics, treatment types, or socioeconomic groups), the model may not perform well in more diverse or less represented scenarios. Ultimately, the default parameters we have chosen for Random Forest Feature selection due to superior performance than grid-search method may not be applicable or effective for different datasets.

### Comparison with Previous Studies

The findings that communication around risk, social behaviors, and empathy play a crucial role in influencing donor behavior have been documented in a previous study^29^, where it was found that emotionally compelling narratives significantly improve crowdfunding outcomes. Research highlighting the financial burden^30^ of cancer care provides further support for income loss as a significant predictor. The importance of medical conditions like being in a hospital or receiving chemotherapy treatment is supported by studies showing that campaigns related to severe or high-cost treatments, such as advanced-stage cancers, often receive more attention and donations^31^. Our study also sheds light on new insight on medical crowdfunding. The analysis introduces the importance of specific linguistic features, such as the use of positive language and social behavior indicators, which were not as thoroughly explored in earlier studies. The involvement of healthcare organizations as a significant predictor suggests that campaigns backed by institutions might have better access to resources and are perceived as more credible.

## Conclusions

The reliance on crowdfunding for conditions associated with high-importance features like hospital stays and income loss highlights potential gaps in healthcare funding. Policymakers need to assess where these gaps exist and consider reforms or additional support mechanisms to reduce the need for crowdfunding in critical areas. The variability in the importance of different features may also point to inequities in access to healthcare and financial support. Policymakers can better target support to vulnerable populations such as those with comorbidities. A crucial policy objective is ensuring that all individuals, regardless of their medical or financial situation, have equal access to medical care without crowdfunding.

## Data Availability

The data and models used to support the findings of this study are available from the corresponding author upon request.

## Conflicts of Interest

The authors declare that there is no conflict of interest regarding the publication of this paper.

## Acknowledgements

The authors thank the Department of Psychiatry and Cancer Center at the College of Medicine, University of Illinois Chicago, for supporting computing resources and research, and J.G., E.M. and R.C. for their valuable feedback.

## Contributions

R.B., J.G., and E.M. conceptualized the problem. R.B., V.S., and A.R. wrote the code and generated tables and figures. L.B. contributed to the data collection, coding and analysis. R.B. and A.R. wrote the manuscript.

## AI Disclosure

The authors acknowledge the use of AI tools for editing the manuscript.

## Notes

### Competing Interest Statement

The authors have declared no competing interest.

### Funding Statement

This study did not receive any funding

### Summary of Updates

This version of the manuscript has been revised to update the authors' order.

