## supplementary tables for "Robust cancer crowdfunding predictions: Leveraging large language models and machine learning for success analysis" (from another version of this preprint: version 1, 2025-02-25)

**Table1: Linguistic Predictors**

| Analytic Thinking | Percentage of words suggesting formal, logical, and hierarchical thinking patterns |
| --- | --- |
| Clout | Percentage capturing relative social status, confidence, or leadership in writing or talking |
| Authenticity | Percentage indicating how much people reveal themselves in an authentic or honest manner |
| Tone | Percentage reflecting the overall emotional tone of the text |
| Cognition | Percentage of words that reflect cognitive processes such as thinking, knowing, and reasoning |
| Politeness | Percentage indicating the degree of politeness in the text |
| Empathy | Percentage of words expressing affection and empathy |
| Social Behavior | Percentage of words related to social interactions and behaviors |
| Prosocial | Percentage of words reflecting prosocial behaviors and attitudes |
| Linguistic Content | Percentage capturing the richness of linguistic content in the text |
| Communication | Percentage reflecting the effectiveness and clarity of communication in the text |
| Family Specific | Percentage of words specifically mentioning family members or related topics |
| Friend Specific | Percentage of words specifically mentioning friends or related topics |
| Lifestyle | Percentage of words related to lifestyles and daily living |
| Physical Behavior | Percentage of words referring to physical activities and behaviors |
| Religion | Percentage of words related to religious topics and spirituality |
| Reward | Percentage of words mentioning rewards and positive outcomes |
| Risk | Percentage of words mentioning risks and potential negative outcomes |
| Past Tense | Percentage of words in the text referring to past events |
| Present Tense | Percentage of words in the text referring to present events |
| Future Tense | Percentage of words in the text referring to future events |

**Table 2: Medical, social and demographic predictors**

| **Category** | **Factors** |
| --- | --- |
| **Medical and Health** | Cancer site, stage or grade of cancer, type of therapy, treatment status, treatment duration, engagement in healthcare activities, comorbidities, association with healthcare organizations, facilities (hospices, hospitals, nursing homes, skilled nursing facilities, emergency rooms) |
| **Emotional States** | Stress, anxiety, mental or behavioral dysfunctions, chronic diseases, cognitive content of communications, expression of social concerns, informal language (swear words, online abbreviations), sentiment (positive, negative, neutral), trust in medical care facilities |
| **Employment and Education** | Employment status, work disruption (for individuals 18 and older), school absenteeism, work absenteeism, parent's work disruption (for beneficiaries under 18) |
| **Financial and Medical Support** | Lack of sick leave, lack of medical attention, income loss, family income loss, struggles with medical expenses, housing expenses, food expenses, transportation expenses |
| **Medical Procedures and Treatments** | Laboratory procedures, diagnostic procedures, cancer-specific treatments (chemotherapy, radiotherapy), preventive procedures, alternative physical therapy, occupational therapy, mental health or substance abuse therapy |
| **Organizations and Facilities** | Association with healthcare-related organizations, professional societies, self-help organizations, relief organizations, receiving care in specific facilities (hospices, hospitals, nursing homes, skilled nursing facilities, emergency rooms, mental substance abuse facilities) |
| **Mental State** | Presence of stress, anxiety, mental or behavioral dysfunctions, chronic diseases |
| **Sentiment** | Sentiment analysis (positive, negative, neutral) |
| **Trust** | Level of trust in medical care facilities |
